# Open Science Practices Among Authors Published in Complementary, Alternative, and Integrative Medicine Journals: An International, Cross-Sectional Survey

**DOI:** 10.1101/2024.02.26.24303402

**Authors:** Jeremy Y Ng, Brenda Lin, Liliane Kreuder, Holger Cramer, David Moher

**Affiliations:** Centre for Journalology, Ottawa Methods Centre, Ottawa Hospital Research Institute, Ottawa, Ontario, Canada; Institute of General Practice and Interprofessional Care, University Hospital Tübingen, Tübingen, Germany; Robert Bosch Center for Integrative Medicine and Health, Bosch Health Campus, Stuttgart, Germany; School of Epidemiology and Public Health, University of Ottawa, Ottawa, Canada

**Keywords:** barriers, complementary and alternative medicine, integrative medicine, open science, open science practices

## Abstract

**Background:** Open science practices aim to increase transparency in research and increase research availability through open data, open access platforms, and public access. Due to the increasing popularity of complementary, alternative, and integrative medicine (CAIM) research, our study aims to explore current open science practices and perceived barriers among CAIM researchers in their own respective research articles.

**Methods:** We conducted an international cross-sectional online survey that was sent to authors that published articles in MEDLINE-indexed journals categorized under the broad subject of “Complementary Therapies” or articles indexed under the MeSH term “Complementary Therapies”. Articles were extracted to obtain the names and emails of all corresponding authors. 8,786 researchers were emailed our survey, which included questions regarding participants’ familiarity with open science practices, their open science practices, and perceived barriers to open science in CAIM with respect to participants’ most recently published article. Basic descriptive statistics was generated based on the quantitative data.

**Results:** The survey was completed by 292 participants (3.32% response rate). Results indicate that the majority of participants were “very familiar” (n = 83, 31.68%) or “moderately familiar” (n = 83, 31.68%) with the concept of open science practices while creating their study. Open access publishing was the most familiar to participants, with 51.96% (n = 136) of survey respondents publishing with open access. Despite participants being familiar with other open science practices, the actual implementation of these practices was low. Common barriers participants experienced in implementing open science practices include not knowing where to share the study materials, where to share the data, or not knowing how to make a preprint.

**Conclusions:** Although participants responded that they were familiar with the concept of open science practices, the actual implementation and uses of these practices were low. Barriers included a lack of overall knowledge about open science, and an overall lack of funding or institutional support. Future efforts should aim to explore how to implement methods to improve open science training for CAIM researchers.

## Background

In an era where knowledge is abundant but not necessarily available, the emerging open science movement is a critical step towards accessible and transparent science. Open science aims to increase research availability and reach across different audiences through open data, open access platforms, transparent peer review methods, and public access [1–6]. Specifically, the movement seeks to increase the amount of science research that is accurately being disseminated to the public as well as trust in science. Prior to the open science movement, access to research data was more limited, had inadequate reporting guidelines, and lacked reproducibility [7,8].

Many measures have been taken to increase study data availability, allowing for improved data assessment and transparency [8–10]. For example, tools such as the Open Science Framework have been created for use as a centralized platform for open workflow throughout the research process [2,11]. These measures allow researchers to better understand, modify, verify, and replicate study data [2,12,13]. In addition, open data encourages the reuse of existing data to avoid redundancy in scientific literature [14]. A crucial part of open science includes open access publishing, which enables research to be more freely communicated with the public. The term ‘open access’ encompasses different materials such as preprints, open access journals, and post-prints, all of which allow more equitable access to scientific information [9,10,15]. Although ‘closed access’ journals still exist, many have now adapted to varying degrees of open access publishing models [10].

For example, a common publishing model is hybrid open access: a model where authors can make their specific articles free for anyone to access through a payment to the publisher, while maintaining a subscription model for the rest of the journal [16]. Despite movement towards an open science landscape, many barriers still obstruct its full implementation. Challenges include a lack of formal education procedures for teaching open science, low incentive to practice open science, and increased publishing costs [2,8,17].

### Open Science Research in Complementary, Alternative, and Integrative Medicine

Little work has previously been conducted at the intersection of open science practices and complementary, alternative, and integrative medicine (CAIM). Therefore, investigating this intersection may offers insight into the current state of open science practices among CAIM research(ers). While the primary language surrounding CAIM have previously been focused on the words ‘alternative’, and ‘complementary’, ‘integrative medicine’ has since become essential to the discourse around CAIM. The word ‘alternative’ typically is focused on therapies that are not used in conventional medicine, whereas ‘complementary’ medicine combines a non-mainstream approach with conventional medicine. Lastly, ‘integrative’ medicine considers the usage of both conventional and unconventional approaches in a coordinated, multimodal manner to provide a more holistic form of care [18–20]. For the purpose of this study, all of the terms described will be collectively referred to as CAIM. It is challenging to determine what therapies are considered to be CAIM due to changes in time periods and geographical locations which may consider traditional forms of medicine to be conventional care [19]. To date, the most updated operational definition of CAIM encompasses 1561 CAIM terms, including 604 unique medications and therapies such as acupuncture and micronutrient supplements [19]. Some challenges associated with implementing CAIM therapy in mainstream healthcare and clinical practice include patient acceptance and understanding of provided CAIM treatments, adequate regulation, and evidence of efficacy [21,22]. Within academic publishing, open science practices surrounding CAIM research are not well defined. To the best of our knowledge, our research on the use of open science practices for CAIM is the first study of its kind. Open science practices have been previously researched in other fields including economics, psychology, and pharmacology through participation in surveys [23–25]. Data from these surveys have been foundational in determining where to focus further efforts to improve open science components in these respective fields of research.

In this study, a cross-sectional online survey methodology was used to determine what open science practices are used by CAIM researchers in their most recently published research article. The objective of this study was to explore the usage of open science practices among CAIM researchers by asking questions regarding their open science practices, such as open access, posting a preprint, and utilizing a reporting guideline, as well as to determine what barriers prevented participants from engaging in these open science practices.

## Methods

### Open Science Statement

This study protocol was registered on the Open Science Framework (OSF) prior to any data collection and can be found here: https://doi.org/10.17605/OSF.IO/P9V8F. All the study materials, such as de-identified survey responses can also be found on OSF: https://doi.org/10.17605/OSF.IO/5JXAH. Ethics approval was granted by the Ottawa Health Science Network (OHSN) Research Ethics Board (REB number: 20230376-01H). This manuscript is reported in accordance with the STROBE (Strengthening the Reporting of Observational Studies in Epidemiology) cross-sectional design reporting guideline [29] as well as using the CHERRIES checklist (Checklist for Reporting Results of Internet E-Surveys) [30].

### Sampling Framework

An international, cross-sectional survey was designed to determine if CAIM researchers apply open science practices specifically to their own published studies indexed in MEDLINE. A single search strategy was conducted by BL on OVID MEDLINE to identify articles published between the dates of January 1, 2018, and May 1, 2023, in MEDLINE-indexed journals categorized under the broad subject of “Complementary Therapies” [26] or articles indexed under the MeSH term “Complementary Therapies” **(Table 1).** This search strategy was used as it is reasonable to assume that a large proportion of these authors identify as CAIM researchers. This search strategy excluded all non-research articles (e.g., editorials, opinions, commentaries, etc.) via a search strategy line. Additionally, records were manually cleaned by BL and LK after the search results were exported to remove any residual non-research records, any duplicate name and email address pairs, as well as to identify authors’ most recently published research articles. All authors who have published in the journals and fit the criteria detailed in the sampling framework were selected to take part in our anonymous, online survey.

**Table 1:** OVID Medline Search Strategy for Author Name and Email Address Retrieval.

|  |
| --- |
| **Database: Ovid MEDLINE(R) ALL <1946 to May 17, 2023>**<br>**Search Strategy:**<br>**1** ("0014162" or "0367526" or "100886202" or "100888586" or "100958201" or "100965420" or "101088661" or "101140517" or "101181180" or "101199657" or "101225531" or "101232990" or "101233160" or "101269884" or "101313855" or "101476585" or "101490763" or "101504416" or "101518405" or "101551481" or "101556804" or "101603118" or "101698453" or "101719675" or "101761232" or 16930290R or "7511141" or "7610364" or "7901431" or "8207427" or "8211546" or "8303080" or "8507710" or "8600658" or "8904486" or "8913656" or "9112559" or "9211576" or "9304117" or "9308777" or "9426370" or "9438794" or "9502013" or "9506953" or "9508124" or "9705340" or "9813115" or "9918283075806676").jc. (114605)<br>**2** exp Complementary Therapies/ (244672)<br>**3** 1 or 2 (317445)<br>**4** limit 3 to (address or autobiography or bibliography or biography or case reports or clinical conference or comment or congress or consensus development conference or consensus development conference, nih or dataset or dictionary or directory or duplicate publication or editorial or electronic supplementary materials or "expression of concern" or festschrift or lecture or legal case or legislation or letter or news or newspaper article or overall or patient education handout or periodical index or personal narrative or portrait or published erratum or "research support, american recovery and reinvestment act" or research support, nih, extramural or research support, nih, intramural or research support, non us gov't or research support, us gov't, non phs or research support, us gov't, phs or retracted publication or "retraction of publication" or technical report or video-audio media or webcast) (131621)<br>**5** 3 not 4 (185824)<br>**6** limit 3 to (english language and dt=20180101-20230501) (48531) |

### Participant Recruitment

Only researchers who were identified using our sampling framework were contacted by JYN to take part in the study. The prospective participants were contacted via email to fill out the survey voluntarily and anonymously. The emails were distributed to the authors in our sample using Microsoft Outlook Mail Merge software [27]. The email provided details about the study goals and included a link to an informed consent form, to which participants agreed before accessing the online survey. An initial screening question was also included in the beginning of the survey, which resulted in only participants that published their research article on a topic specific to one or more CAIM medicine therapies to be eligible to complete the survey. Participants were given a total of four weeks to participate in the survey, with email reminders being sent after the first, second, and third weeks, respectively, following the original invitation email. The survey was open from October 4, 2023 until November 15, 2023. Survey participation was voluntary; no financial compensation to complete the survey was offered. Apart from the screening question, participants were able to skip any other survey questions they did not wish to answer. Participants were also able to use a ‘Back’ button to change their answers, however, participants were not able to withdraw their responses once the survey was submitted as their identities were not collected.

### Survey Design

The survey was designed using the University of Ottawa’s approved version of Survey Monkey software. Each survey was personalized for individual participants using Survey Monkey’s custom variable logic feature [28] to specify the title and the Digital Object Identifier (DOI) of their study within the survey questions. This personalization allowed for the creation of unique Survey Monkey links for each participant that was distributed using Microsoft Outlook Mail Merge software [27]. The survey was initially piloted amongst the authors to ensure that the survey design was ready to be widely distributed. Each author received a survey invitation via email and a link to the personalized survey on Survey Monkey which contained an implied consent agreement. The survey consisted of 31 multiple-choice questions. It began with 7 preliminary questions regarding general demographic information. Five questions were then asked regarding research background. Following these demographics questions, the next 17 questions asked participants about their familiarity and usage of open science practices prior to or after starting the research study as listed in their survey invitation. Lastly, survey respondents were given the option to write anything else they wanted to share at the end of the survey.

### Data Analysis

This study had no formal hypotheses. Basic descriptive statistics including frequencies and percentages was generated based on the analysis of the quantitative data. A thematic content analysis was also conducted on a qualitative survey question. BL and LK team members coded the survey responses. To reach an agreement on their respective codes and create a coding framework, multiple rounds of discussion took place about deductive and inductive coding, and coding category exclusivity and specificity.

## Results

### Demographics

Out of 11 476 emails that were sent, there was a total of 2694 emails that bounced back, and therefore was not included to calculate the total response rate. Overall, a total of 292 respondents participated in our survey (3.32% response rate; 100% completion rate). Participants were not required to respond to each question, and so the denominators used to calculate participant responses differed. The survey was considered to be incomplete if no questions were answered after the initial screening question at the beginning of the survey. Participants spent an average of 9 minutes and 39 seconds to complete the survey. As summarized in **Table 2** demonstrating participant demographics, slightly over half of the participants considered CAIM research to be their primary research interest (n = 155), with the remainder considered CAIM to be a secondary research interest. The majority of participants were located in North America (n = 77, 27.40%), Asia (n = 72, 25.62%), or Europe (n = 68, 24.20%), with the rest located in Africa (n = 19; 6.76%), South America (n = 21, 7.47%), and Australasia (n = 24, 8.54%). The majority of participants had completed a doctoral level of education (n = 201, 71.53%), and currently have positions mainly as a faculty member or principal investigator (n = 151, 54.91%) or clinician (n = 34, 12.36%). The majority of participant’s primary research area was identified to be clinical research (n = 143, 51.81%). Raw survey responses (personal identifiers redacted) can be found on OSF: https://osf.io/qu5xr.

**Table 2:** Characteristics of Survey Participants.

|  |  |
| --- | --- |
| Sex (n = 140) | n (%) |
| Female | 140 (49.65%) |
| Male | 138 (48.94%) |
| Non-Binary | 0 (0.00%) |
| Prefer not to say | 1 (0.35%) |
| Prefer to self-describe | 3 (1.06%) |
| Age (n = 282) |  |
| 18-24 | 0 (0.00%) |
| 23-34 | 36 (12.77%) |
| 35-44 | 86 (30.50%) |
| 45-54 | 76 (26.95%) |
| 55-64 | 45 (15.96%) |
| 65 or older | 34 (12.06%) |
| Prefer not to say | 5 (1.77%) |
| All levels of education earned (n = 281) |  |
| High school | 132 (46.98%) |
| Undergraduate degree (BSc, BA, BEng, BScN, etc.) | 122 (43.42%) |
| Professional undergraduate degree (MD, MBSS, DDS, PharmD, etc.) | 78 (27.76%) |
| Master's degree (MSc, MA, MPH, MLIS, etc.) | 127 (45.20%) |
| Doctoral degree (PhD, EdD, etc.) | 201 (71.53%) |
| Postdoctoral fellow | 78 (27.76%) |
| CAIM practitioner (e.g., naturopath, homeopath, chiropractor, etc.) | 41 (14.59%) |
| Other | 26 (9.25%) |
| Visible minority (n = 281) |  |
| Yes | 44 (15.66%) |
| No | 224 (79.72%) |
| Prefer not to say | 13 (4.63%) |
| Disability (n = 278) |  |
| Yes | 8 (2.88%) |
| No | 265 (95.32%) |
| Prefer not to say | 5 (1.80%) |
| Caregiver (n = 281) |  |
| Yes | 116 (41.28%) |
| No | 158 (56.23%) |
| Prefer not to say | 7 (2.49%) |
| Location (n = 281) |  |
| Africa | 19 (6.76%) |
| Asia | 72 (25.62%) |
| Australasia | 24 (8.54%) |
| Europe | 68 (24.20%) |
| North America | 77 (27.40%) |
| South America | 21 (7.47%) |
| Current position (n = 275) |  |
| Graduate student | 12 (4.36%) |
| Postdoctoral fellow | 22 (8.00%) |
| Faculty member/Principal investigator | 151 (54.91%) |
| Research support staff (e.g., research manager, research associate, technician, etc.) | 11 (4.00%) |
| Scientist in industry | 7 (2.55%) |
| Scientist in third sector (e.g., NGO, non-profit) | 5 (1.82%) |
| Government scientist | 8 (2.91%) |
| Clinician | 34 (12.36%) |
| Other | 25 (9.09%) |
| Primary research area (n = 276) |  |
| Clinical research | 143 (51.81%) |
| Preclinical research – in vivo | 19 (6.88%) |
| Preclinical research – in vitro | 23 (8.33%) |
| Health systems research | 30 (10.87%) |
| Methods research | 16 (5.80%) |
| Epidemiological research | 14 (5.07%) |
| Other | 31 (11.23%) |
| CAIM research priority (n = 274) |  |
| Primary | 155 (56.57%) |
| Secondary | 119 (43.43%) |
| Area of CAIM research (n = 273) |  |
| Mind-body therapies | 71 (26.01%) |
| Biologically based practices | 76 (27.84%) |
| Manipulative and body-based practices | 69 (25.27%) |
| Biofield therapy | 11 (4.03%) |
| Whole medical systems | 75 (27.47%) |
| Other | 51 (18.68%) |

### Familiarity with Open Science Practices

When asked about participants’ familiarity with open science practices when conducting their study, the majority of participants were either “very familiar” (n = 83, 31.68%) or “moderately familiar” (n = 83, 31.68%) with open science practices. Only a small percentage of participants (6.87%, n = 18) reported being not familiar.

Upon further exploration, participants were asked about their familiarity with various open science practices when writing their study. The majority of participants expressed being “very familiar” with the practice of open access (n = 173, 65.53%). Only 2.64% of respondents (n = 7) claimed to not be familiar with open access. Additionally, participants were also “very familiar” with the open science practices of preprints (n = 106, 40.93%), registration (n = 112, 43.08%), and reporting guidelines (n = 124, 47.69%). Respondents were relatively evenly divided between “very familiar” or “moderately familiar” for the practices of open data (n = 81, 31.15% and n = 72, 27.69%, respectively), and participant and public involvement (n = 77, 29.73%, and n = 69, 26.64%, respectively). Open material practices were the least familiar among all open science practices, with participants ranging from being slightly familiar (n = 47, 18.22%), to having only 20.93% (n = 54) of participants being very familiar with the practice. Furthermore, open material had the highest percentage of respondents who claimed to be “not at all familiar” with the practice, with a value of 15.12% (n = 39) **(Figure 1)**.

**Figure 1:**
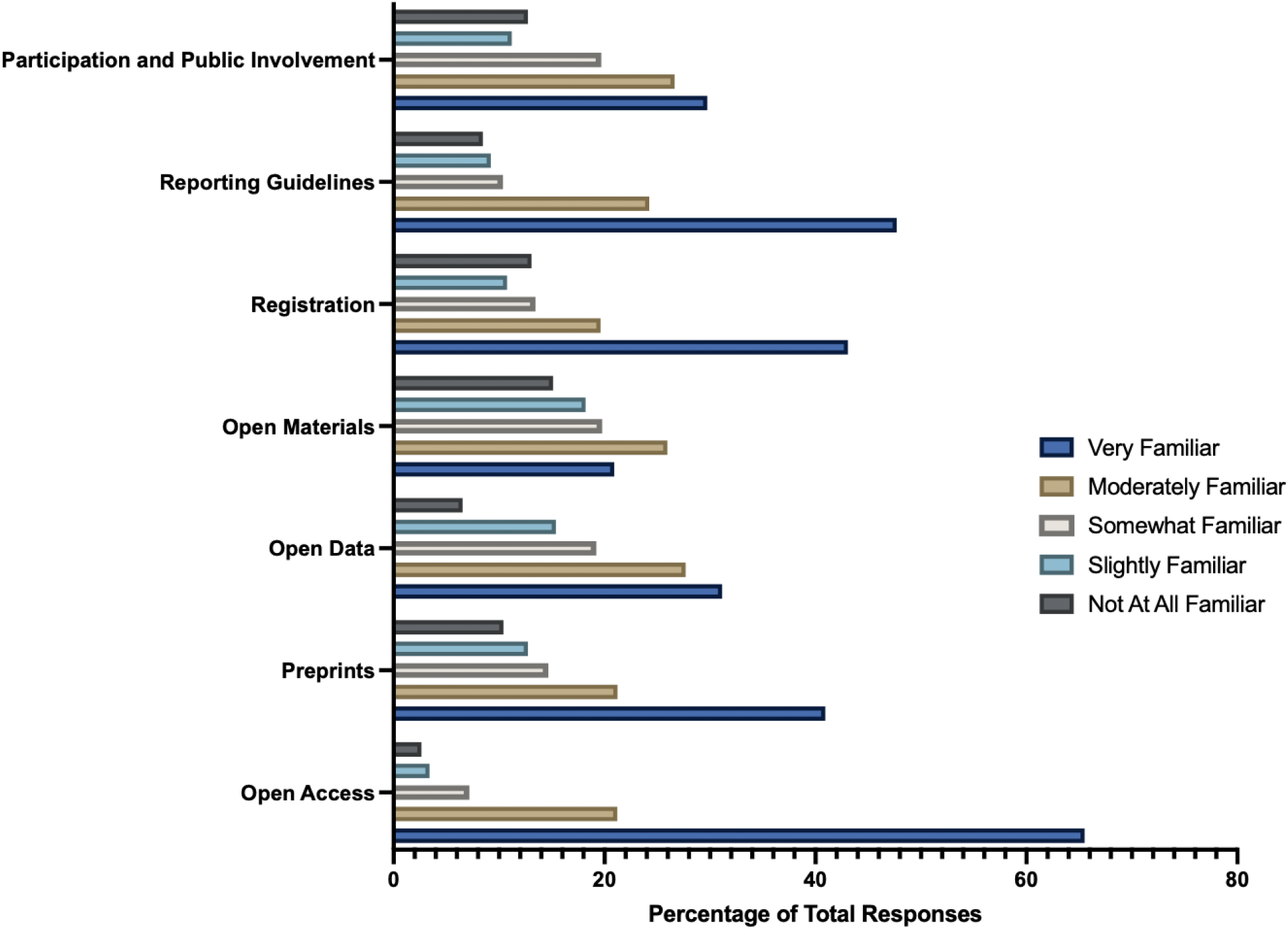
Participant Familiarity with Open Science Practices.

### Incentives in Open Science

When asked about the incentives available before participants began writing their articles, the majority of participants (n = 173, 66.03%) reported having access to additional funding to facilitate open science practices. This funding included support for open access charges and resources for employing additional staff to assist in preparing data for open sharing.

Other incentives provided to participants included clearer communication as to why open science is valuable for research (n = 116, 44.27%), practical support from their institution to conduct open science (n = 122, 46.56%), and additional training on how to perform open science practices (n = 100, 38.17%). Notably, only around 25% of respondents had staff trained in open science practices (n = 68, 25.95%), or had a way to get recognized for their performance of open science practices when being hired, promoted, or tenured (n = 70, 26.72%) **(Figure 2)**.

**Figure 2:**
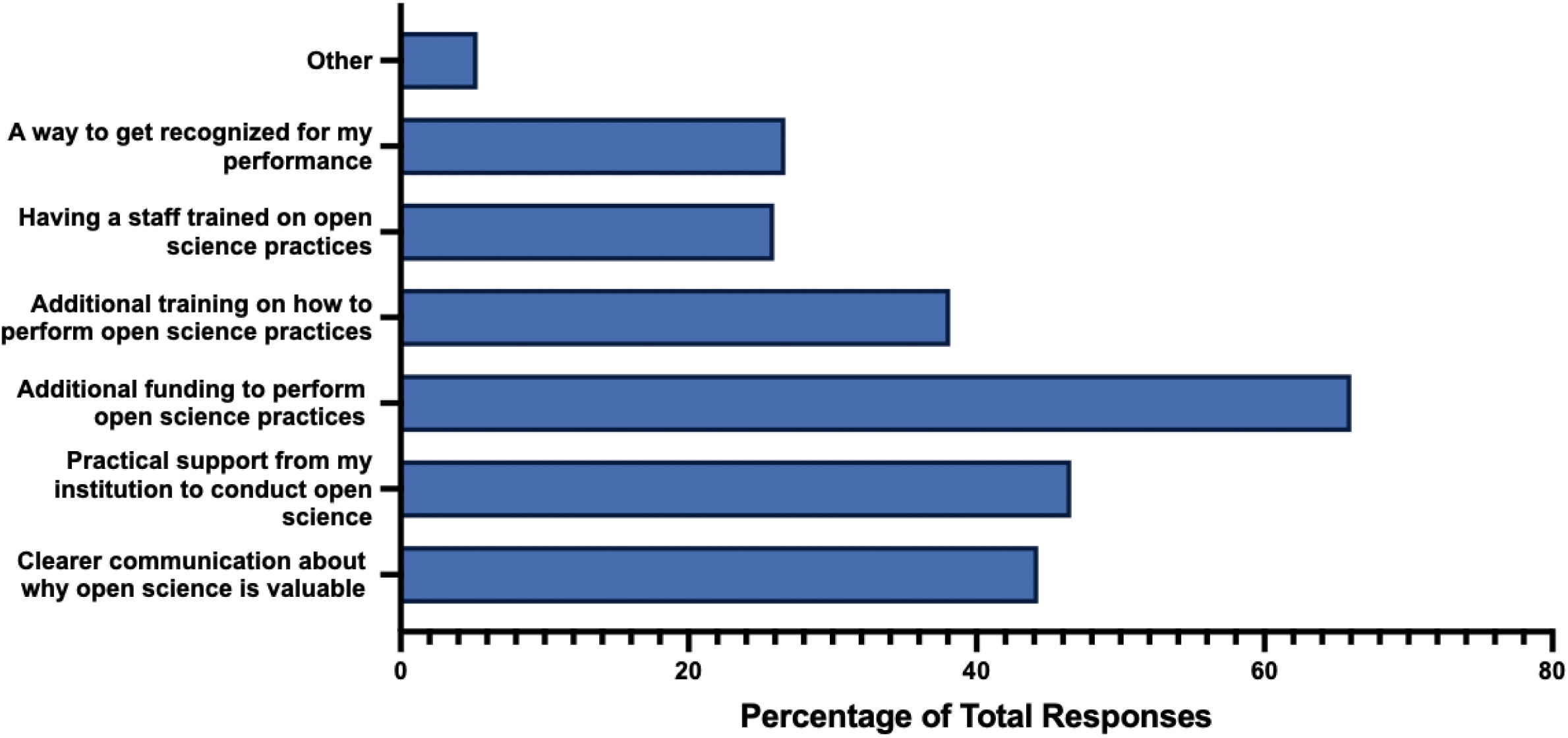
Incentives Available that Would Result in Applying More Open Science Practices.

### Open Access

Roughly half of the participants published their articles open access (n = 136, 51.96%), while a small percentage (n = 26, 9.92%) did not know if their study was published with open access. In exploring barriers related to publishing the study mentioned in the invitation email, the primary obstacle to open-access publishing was a lack of funding support to cover the article processing charges typically associated with open-access journals (n = 161, 64.14%). Other challenges included a lack of knowledge on how to self-archive a paper to make it open access (n = 28, 11.16%), journals not adopting an open-access publishing model (n = 20, 7.97%), or the perception that their institution does not prioritize open-access publishing (n = 18, 7.17%). A small percentage of participants did not see the benefit of making an article open access (n = 8, 3.19%), and approximately a quarter of participants (n = 58, 23.11%) did not perceive any of the mentioned barriers as issues for publishing open access **(Figure 3)**.

**Figure 3:**
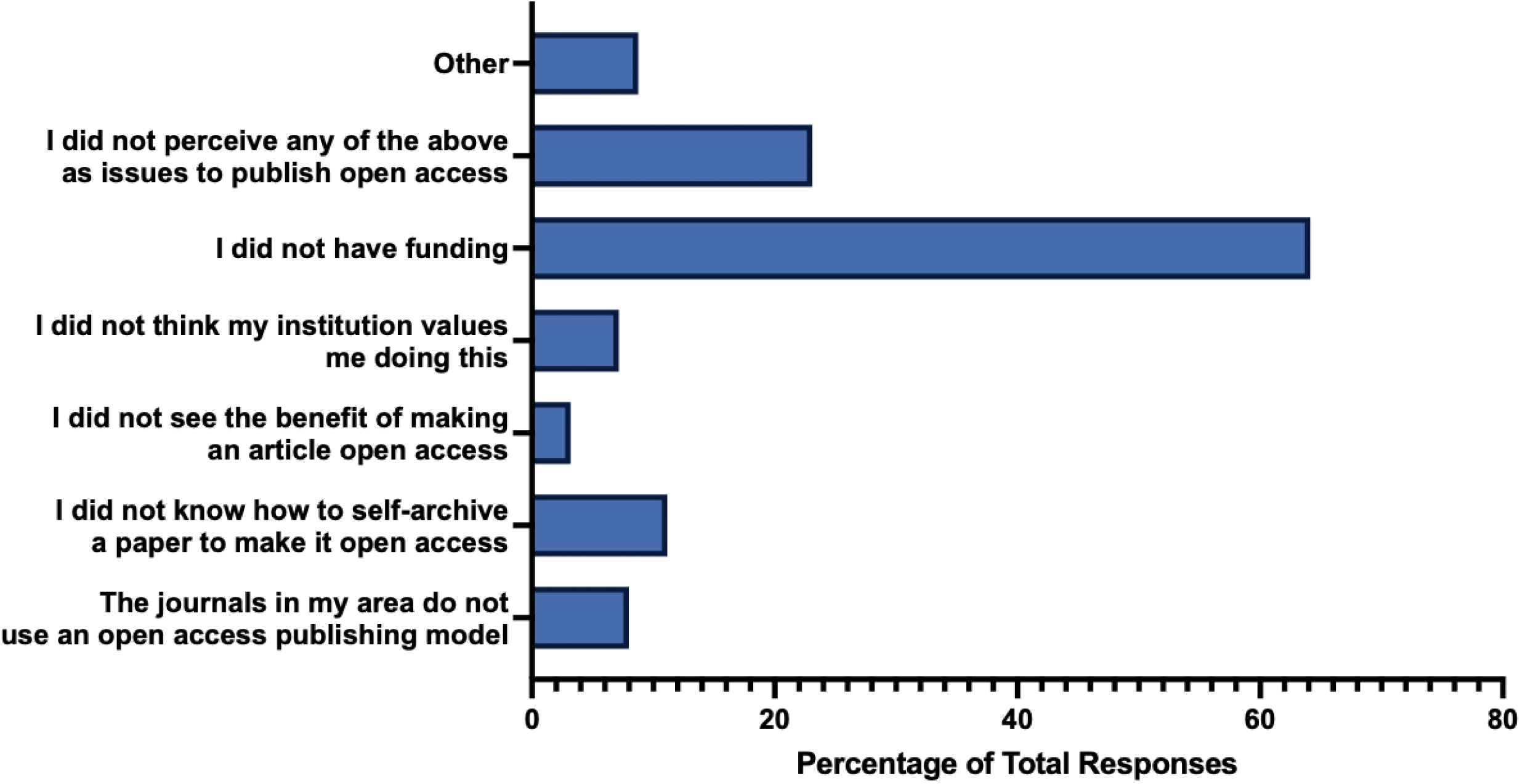
Barriers for Participants to Publish with Open Access.

### Preprinting

When asked if participants created and posted a preprint of their article prior to publishing, the majority of participants answered “no” (n = 204, 79.07%). A smaller percentage answered “yes” (n = 29, 11.24%), while some participants were unsure (n = 25, 9.69%). Regarding the barriers participants faced in creating and posting a preprint of their article, the most common response was a perceived lack of benefit in posting a preprint (n = 81, 32.53%). Another obstacle was a lack of knowledge on how to create a preprint (n = 74, 29.72%). Additionally, approximately a quarter of participants expressed concerns about potential harms associated with sharing work that has not undergone peer review (n = 63, 25.30%), or the belief that creating a preprint might reduce the chances of the work being accepted by a peer-reviewed journal (n = 54, 21.69%). A small percentage of participants mentioned that their institution does not value the creation of preprints (n = 27, 10.84%), or that their institution has an internal process for posting preprints that makes the procedure very time-consuming (n = 5, 2.01%) **(Figure 4)**.

**Figure 4:**
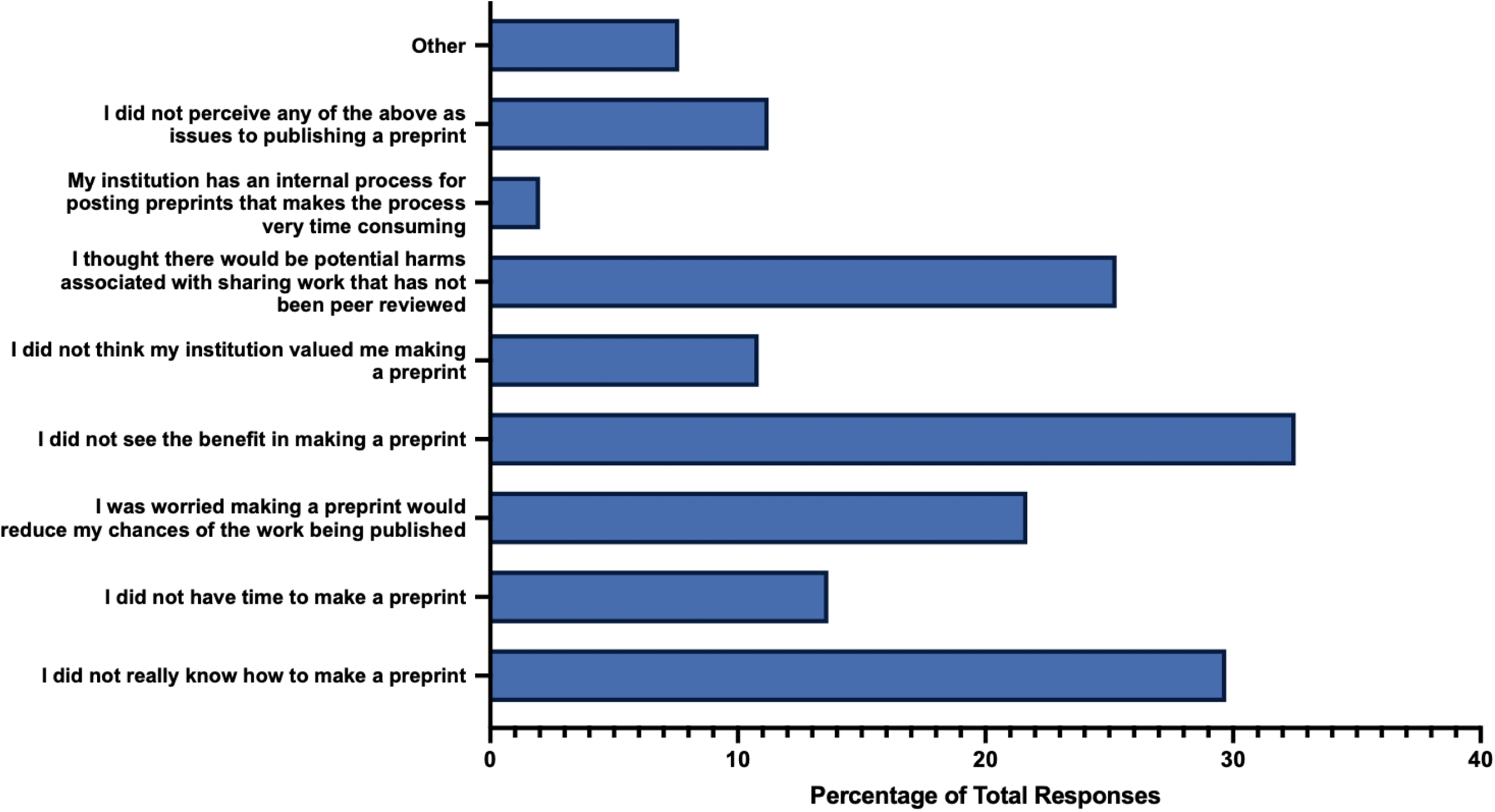
Barriers for Participants to Create and Post a Preprint of Their Respective Study.

### Sharing Data

When asked if participants shared the raw data associated with their study, the majority of participants responded “no” (n = 178, 70.08%), with 23.23% (n = 59) responding “yes”, and 6.69% (n = 17) of respondents being “unsure”. Regarding the barriers participants faced in sharing the raw data when publishing their studies, a significant number indicated they did not know how to prepare data for sharing (n = 54, 21.95%), lacked the time for data preparation (n = 47, 19.11%), or were unsure where to share their data (n = 43, 17.48%). Some researchers refrained from sharing raw data due to restrictions specified in their research consent forms (n = 44, 17.89%), or concerns about patient privacy (n = 39, 15.85%). Some participants had concerns about intellectual property control (n = 41, 16.67%) and the unintended use of secondary data (n = 40, 16.26%). A smaller proportion of researchers expressed concerns about being scooped (n = 16, 6.50%), or that others might discover errors in the data (n = 9, 3.66%). It’s noteworthy that the largest proportion of participants (n = 59, 23.98%) did not perceive any of the mentioned issues as barriers to sharing data **(Figure 5)**.

**Figure 5:**
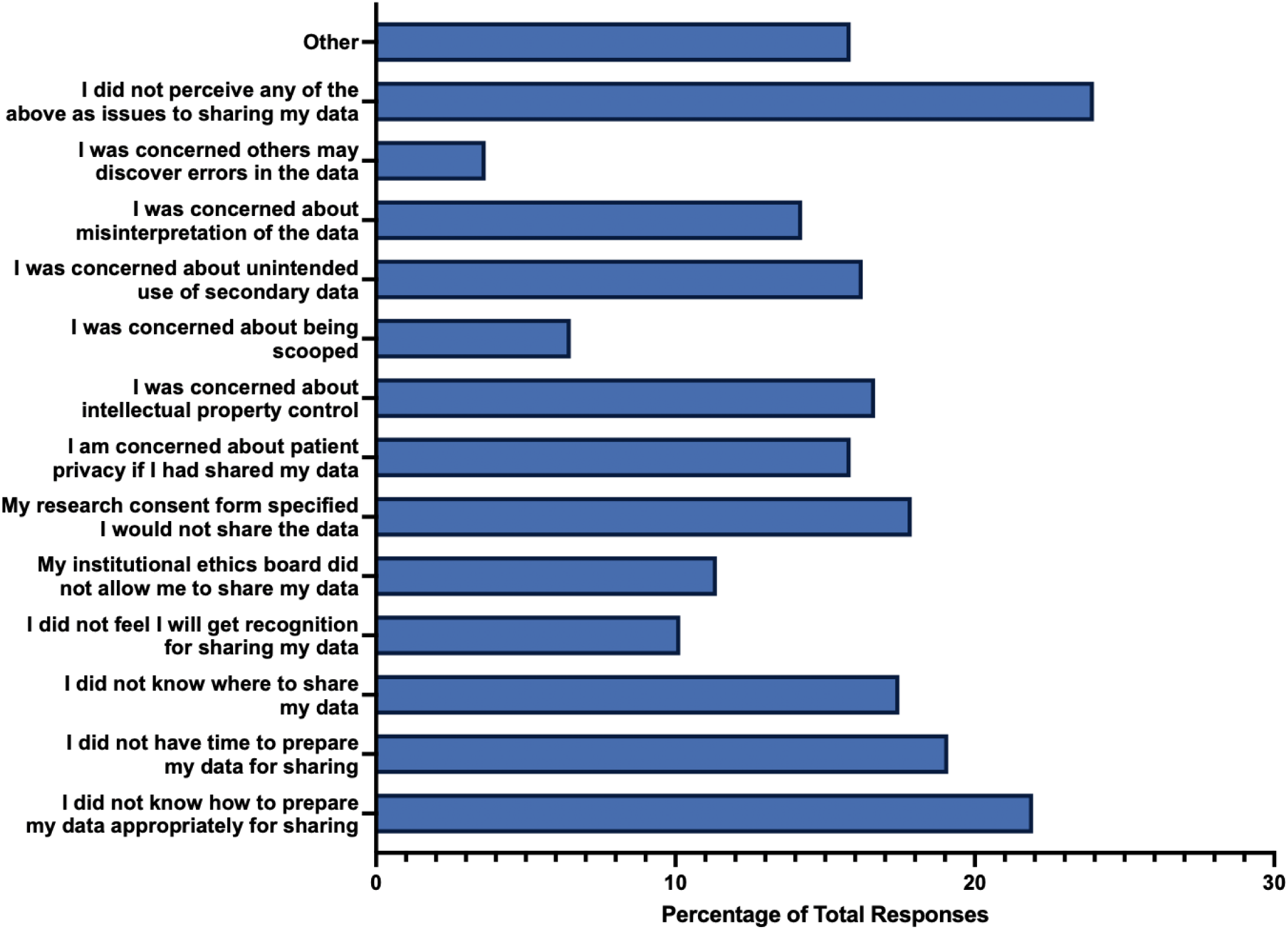
Barriers for Participants to Share the Raw Data of Their Respective Study.

### Sharing Study Materials

The majority of participants did not share study materials associated with their study (n = 149, 59.60%), with only 28.00% (n = 70) of respondents answering “yes”, and 12.40% (n = 31) being unsure. In terms of barriers to sharing study materials, the most common obstacle was participants not knowing how to prepare study materials for sharing (n = 49, 20.86%). Other responses included uncertainty about where to share study materials (n = 44, 18.72%), a lack of time for preparation (n = 36, 15.32%), or the perception that there was no value for others in sharing their study materials (n = 33, 14.04%). Barriers less frequently experienced by participants included concerns about being scooped (n = 7, 2.98%), institutional ethics boards disallowing the sharing of study materials (n = 10, 4.26%), or the perceived cost of sharing study materials (n = 11, 4.68%). However, the majority of respondents (n = 67, 28.51%) did not perceive any of the mentioned issues as barriers to sharing study materials **(Figure 6)**.

**Figure 6:**
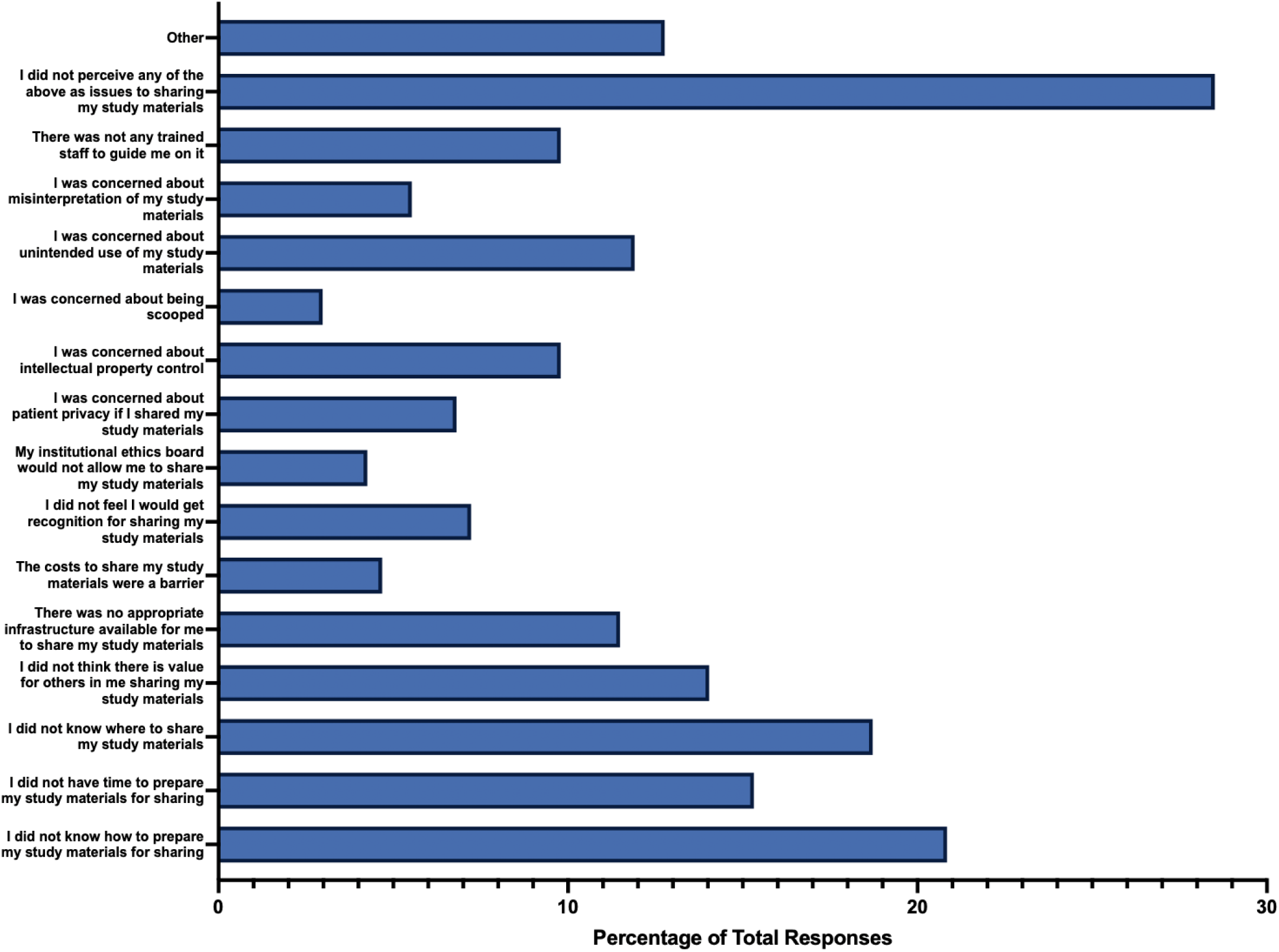
Barriers for Participants to Share the Study Materials of Their Respective Study.

### Registering a Study Protocol

When participants were asked if they registered their study’s protocol, the majority of respondents did not (n = 144, 58.06%). A smaller percentage of respondents answered “yes” (n = 93, 37.50%), and some were unsure (n = 11, 4.44%). Further investigation revealed that the most common barrier participants faced in registering their study protocol was a lack of knowledge about which platform to use for registration (n = 43, 18.53%). The next common barrier was participants not knowing how to create a study registration (n = 40, 17.24%). Other reasons included a perceived lack of time to register the study (n = 26, 11.21%), and the belief that their research area did not lend itself well to registering protocols (n = 26, 11.21%). Smaller barriers included a perception that their institution did not prioritize registering protocols (n = 6, 2.59%), concerns about being scooped if sharing the study plan before publishing (n = 9, 3.88%), or a lack of recognition for taking the time to register the study protocol (n = 10, 4.31%). It’s noteworthy that the largest percentage of respondents (n = 82, 35.34%) did not perceive any of the mentioned issues as obstacles to registering a study protocol **(Figure 7)**.

**Figure 7:**
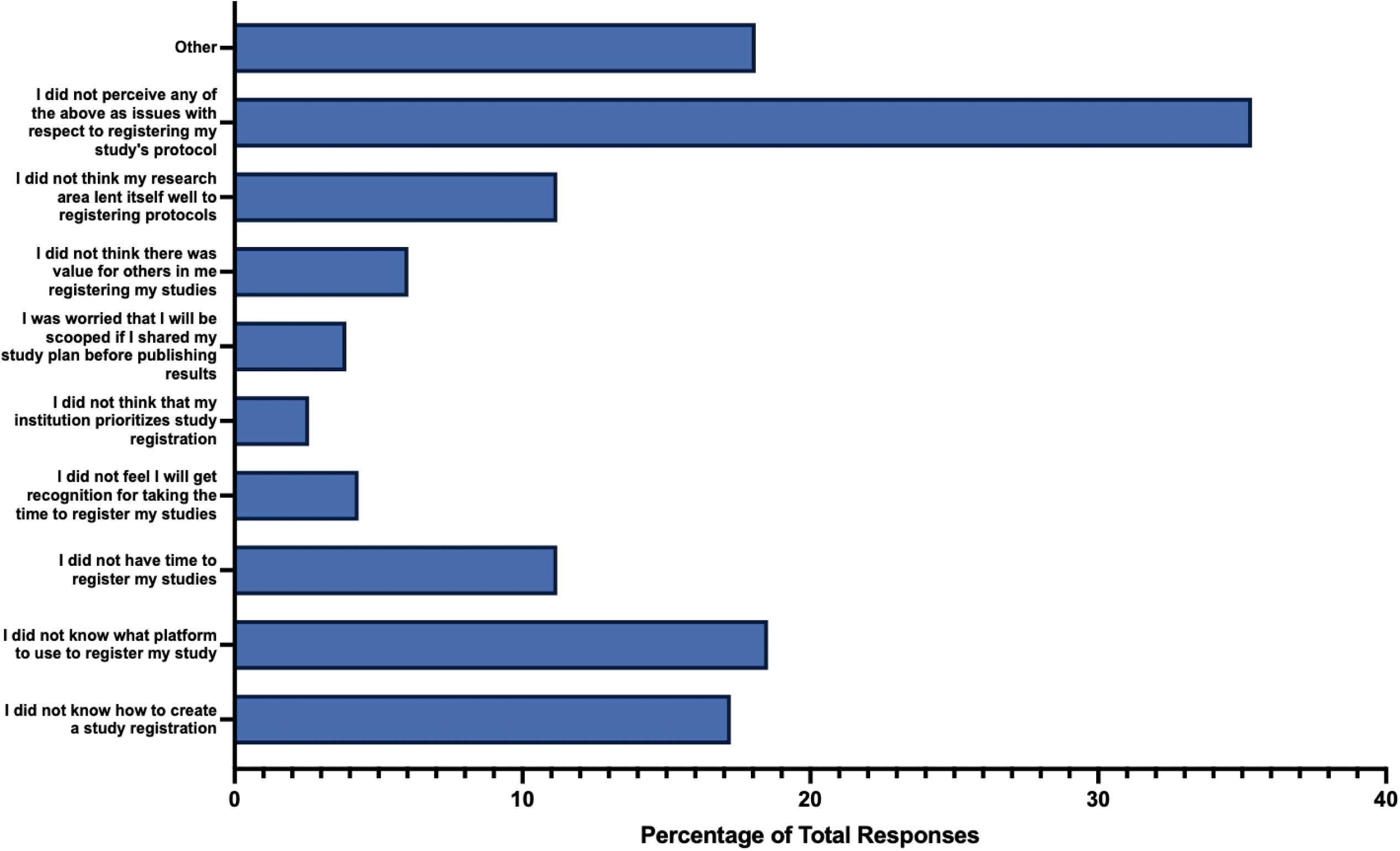
Barriers for Participants to Register their Respective Study Protocol.

### Referencing a Reporting Guideline

When participants were asked if they explicitly used and referenced a reporting guideline checklist in their publication, 57.96% of respondents answered “yes” (n = 142), 33.88% answered “no” (n = 83), and 8.16% did not know (n = 20). Regarding barriers participants faced in using reporting guidelines, the most common obstacle was not knowing where to find the relevant reporting guideline (n = 33, 14.80%), closely followed by the journal not requiring the use of a reporting guideline (n = 27, 12.11%), and not knowing how to use a reporting guideline (n = 23, 10.31%). Additionally, 9.87% of respondents indicated a lack of available reporting guidelines for their type of study (n = 22). A small percentage of respondents felt that they would not receive recognition for taking the time to use a reporting guideline (n = 6, 2.69%) and did not believe that their institution prioritized the use of reporting guidelines (n = 6, 2.69%).

Furthermore, the vast majority of participants did not perceive any of the mentioned issues as barriers to using reporting guidelines (n = 107, 47.89%) **(Figure 8)**.

**Figure 8:**
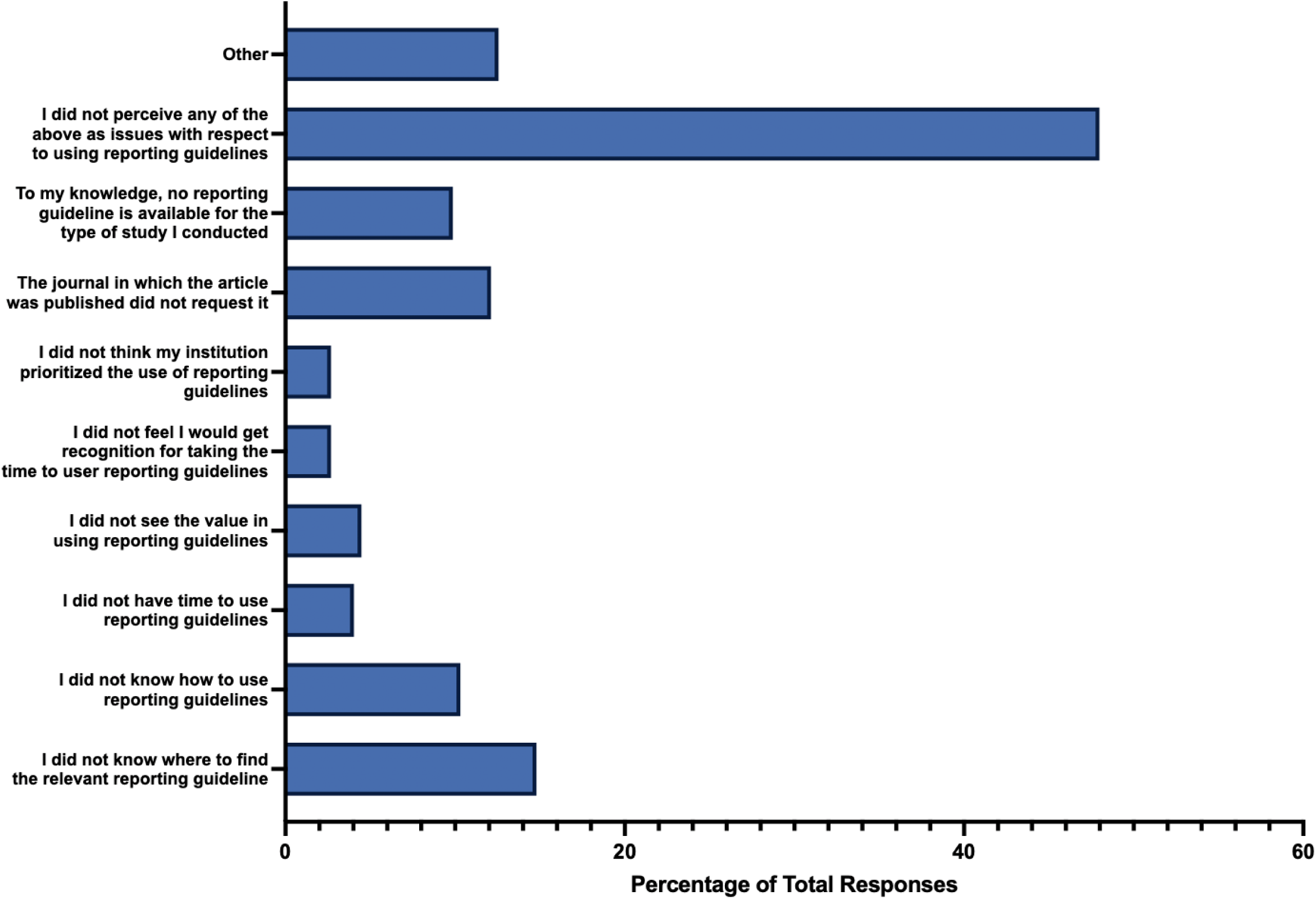
Barriers for Participants to Reference a Reporting Guideline in Their Respective Study.

### Engaging the Public

When asked if respondents engaged participants or members of the public in their study, 60.33% responded “no” (n = 146), 35.54% responded “yes” (n = 86), and 4.13% responded that they did not know (n = 10). Concerning barriers participants faced when engaging participants or members of the public in their study, common challenges included not knowing how to identify individuals from these groups to contribute (n = 42, 18.75%), a lack of understanding on how to incorporate participants or public members in their research (n = 38, 19.69%), and a perceived lack of value in incorporating participants or public members in their research (n = 33, 14.73%). Less common barriers included the belief that they would not be recognized for taking the time to include participants (n = 8, 3.57%), or the perception that their institution did not prioritize incorporating participants (n = 12, 5.36%). Lastly, a substantial proportion of respondents did not perceive any of the mentioned issues as obstacles when engaging participants or members of the public (n = 89, 39.73%) **(Figure 9)**.

**Figure 9:**
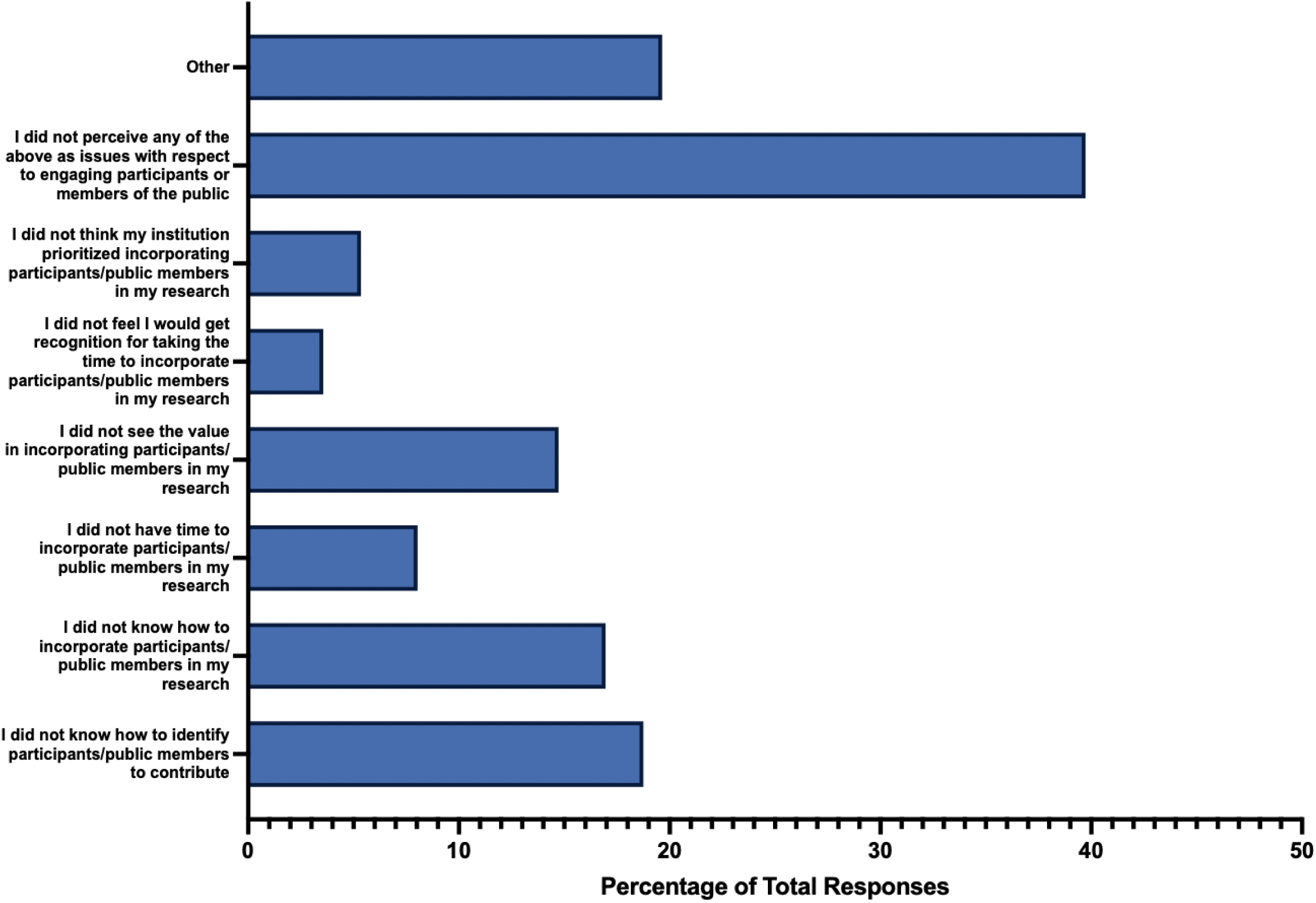
Barriers for Participants to Engage Members of the Public in their Respective Study.

### Thematic Analysis of Open Responses

Thematic analysis of the open responses revealed two major themes: (1) challenges for open science implementation and (2) attitudes towards open science. In the first theme, open responses revealed five sub-themes highlighting different challenges participants encountered and believe were a challenge to open science implementation. Challenges include (1.1) cost of publishing in open access, (1.2) data sharing barriers, (1.3) limited access to open science resources, (1.4) lack of knowledge in open science, and (1.5) a lack of institutional support. The second theme contained two subthemes split into (2.1) positive open science perceptions and (2.2) negative open science perceptions. Positive perceptions included the belief that open science “is the right thing to do” and that open access should be more accessible, while negative open science perceptions included the belief that open access will result in more predatory publishing journals. Coding and thematic analysis data can be found on OSF: https://osf.io/uw8px.

## Discussion

The primary goal of this study was to assess the open science practices of CAIM researchers regarding their respective research articles and identify the barriers they encountered in implementing open science practices. Our findings revealed that while most participants are “very familiar” or “moderately familiar” with the concept of open science, their actual engagement in various open science practices was limited. Participants demonstrated a higher level of familiarity with open access publishing, with approximately half of the respondents indicating that they had published their studies open access. However, despite the reported familiarity with practices such as preprints and open data, participants were less likely to engage in these practices when publishing their own research.

Additionally, our results highlighted an evident knowledge gap in open science practices. A considerable portion of respondents expressed uncertainty or lack of knowledge when it came to preprints, data sharing, study materials sharing, and referencing reporting guidelines. These gaps in knowledge were also evident in the responses about engaging participants or members of the public in research. Open text responses at the end of the survey echoed this sentiment, with many participants citing a lack of training or resources provided by their institutions as hindrances to becoming educated in conducting and consequently engaging in open science practices.

### Comparative Literature

These findings align with prior literature examining open science practices. A current pre-print of a descriptive study conducted by Ng et al. that explored CAIM researchers’ practices and perceived barriers related to open science also identified insufficient funding as a perceived barrier in open access publishing [31]. Similarly, this study found that a lack of knowledge was common for participants to not engage in the open science practice of registering a study protocol or using a reporting guideline, with participants stating that they do not know how to create a study registration, or that they do not know where to find relevant reporting guidelines [31]. This gap in knowledge may be due to a lack of training, since survey respondents appeared to have an overall lack of formal training in open science practices, with the majority of participants having been self-taught [31].

While there are no other comparable studies about open science practices among CAIM researchers, other studies have investigated open science attitudes and practices in other fields. Spitzer et al. investigated factors that either facilitate or prevent researchers in the field of psychology from pre-registering their studies [32]. Their survey found that perceived benefits of pre-registering a study was having increased transparency and a higher quality of research, while major drawbacks to pre-registering that lowered survey respondents’ motivation to pre-register included a loss of flexibility, time costs, and difficulty to justify any changes to or deviations of the study to journal editors or reviewers [32]. Furthermore, the survey similarly found that suggestions to improve preregistration largely included better incentives, such as increased funding, benefits for job applications or tenure, and advantages for publication [32]. In a survey conducted by Ferguson et al. on open science practices of researchers in the field of social sciences including economics, sociology, and psychology, the authors found that the lifetime prevalence of researchers using open science practices has increased from 49% to 87% over the past decade [33]. In particular, the study focused on determining the uses and attitudes of posting code or data, as well as pre-registering hypotheses. The authors assessed how the usage of open science practices have changed over time by asking respondents about their first time they utilized an open science practice, which showed 87% of respondents having used an open science practice by 2020, compared to only 49% by 2010 [33]. In a survey conducted by Christensen et al. on the general attitudes towards the adoption of open science practices across the same research fields, it was found that the growing support for open science is outpacing its adoption, suggesting barriers to open science practice [34]. Similarly, Paret et al. also found that while there is a growing adoption of open science practices among neuroimaging researchers, many did not regularly share their data or preregister their work [35]. The authors found that potential barriers for neuroscientists to preregister their work included the belief that their analyses were too complicated to preregister and that there was not a sufficient reward for preregistration, while barriers to sharing data included challenges to anonymize data and consent forms stating that data would not be shared [35]. Additionally, Houtkoop et al. identified that perceived barriers to data sharing for authors in the field of psychology included a lack of training in sharing data, as well as the belief that data sharing requires additional work [36]. Consistently, challenges such as a lack of support and training emerge as recurrent impediments to the adoption of open science principles.

### Strengths and Limitations

Our study has several notable strengths. Conducting a cross-sectional survey study allows us to collect all the data at one time point which makes this study efficient and cost effective [37]. Our sampling strategy of seeking a relatively large, international participant pool for the survey increases the generalizability of our findings regarding how CAIM researchers apply open science practices. Additionally, the data extraction and cleaning were carried out independently and in duplicate.

With respect to limitations, although we planned to survey a large number of researchers, the response rate was low; furthermore, a proportion of participants likely have changed their affiliation, or otherwise no longer have access to their email, be on vacation or absent, be retired, or have passed away, all of which contributed to nonresponse bias. The survey findings might also be biased towards those who engage in open science practices or those who have reservations or concerns against open science practices, as CAIM researchers with limited interest in open science may be more hesitant or less interested in participating. Additionally, the findings may not be representative of researchers and clinicians who are not English speakers as it is administered in the English language. Lastly, another limitation includes the possibility of participants being unable to accurately recall or omitting details about their open science practices (or lack thereof) they engaged in since months or years have passed since their study was published, leading to recall bias [34].

## Conclusions

In this study, we conducted a cross-sectional online survey to explore the open science practices and associated barriers preventing participants from engaging in these open science practices among CAIM researchers in their most recently published articles. The survey gathered responses from participants regarding their experiences and engagement with open science practices in the realm of CAIM research. The collected data provides valuable insights into the current landscape of open science practices within this field. Our survey revealed a noteworthy disparity between participants’ familiarity with open science concepts, such as open access and pre-printing, and their actual implementation. Although many participants were familiar with these practices, the actual application and usage of open science practices in the respective articles of survey participants was relatively low. Cost emerged as a significant barrier for some participants to participate in the open science practice of open access publishing, consequently affecting their ability to publish in open access journals. Additionally, a consistent theme among respondents was a lack of knowledge in open science practices, acting as a barrier for participants to create and post a preprint, registering a study protocol, or sharing data or study materials. While previous research has suggested strategies to enhance open science implementation in diverse fields, our study uniquely pinpoints specific areas within CAIM research that require increased attention and implementation of open science practices. The identified barriers underscore the need for targeted interventions to address challenges related to cost and the availability of resources and training. We anticipate that our study findings will serve as a valuable resource for researchers and professionals in the field of CAIM, offering insights to improve the adoption of open science practices.

## Data Availability

All data and materials associated with this study have been posted on the Open Science Framework.

https://doi.org/10.17605/OSF.IO/5JXAH

## List of Abbreviations

CAIM: complementary, alternative, and integrative medicine
CHERRIES: checklist for reporting results of internet e-surveys
DOI: digital object identifier
OHSN: Ottawa health science network
OSF: open science framework
REB: research ethics board
STROBE: strengthening the reporting of observational studies in epidemiology

## Declarations

### Ethics Approval and Consent to Participate

We sought and were granted ethics approval by the Ottawa Health Sciences Network Research Ethics Board (REB number: 20230376-01H). All methods were carried out according to this REB’s guidelines. Informed consent was obtained from all survey participants.

### Consent for Publication

Not applicable.

### Availability of Data and Materials

All data and materials associated with this study have been posted on the Open Science Framework and can be found here: https://doi.org/10.17605/OSF.IO/5JXAH

### Authors’ Contributions

JYN: designed and conceptualized the study, collected and analysed data, drafted the manuscript, and gave final approval of the version to be published.

BL: assisted with the collection and analysis of data, drafted the manuscript, made critical revisions to the manuscript, and gave final approval of the version to be published.

LK: assisted with the collection and analysis of data, drafted the manuscript, made critical revisions to the manuscript, and gave final approval of the version to be published.

HC: assisted with the analysis of data, made critical revisions to the manuscript, and gave final approval of the version to be published.

DM: assisted with the design and concept of the study and the analysis of data, made critical revisions to the manuscript, and gave final approval of the version to be published.

## Acknowledgements

We gratefully acknowledge Navila Asgar for her contributions to our study protocol.

